# Clinical outcomes of early aspirin versus non-aspirin NSAID use in adults hospitalized with influenza: A retrospective study

**DOI:** 10.64898/2026.08.05.26359840

**Authors:** Suk Yin Chan-Colenbrander, Qi Wang

## Abstract

Seasonal influenza remains a major cause of morbidity and mortality worldwide. Although neuraminidase inhibitors improve clinical outcomes, influenza-related deaths persist. We evaluated the associations of early aspirin and non-aspirin nonsteroidal anti-inflammatory drug (NSAID) use with clinical outcomes in adults hospitalized with influenza. This retrospective study included adults admitted to the University of Minnesota Medical Center from 2016 to 2018. Multivariable logistic and Cox regression models adjusted for age, sex, race, smoking status, influenza vaccination status, and cardiovascular burden were used to evaluate the associations of early aspirin and NSAID use with clinical outcomes. Among 2,816 patients screened, 320 had laboratory-confirmed influenza. Compared with unvaccinated patients, vaccinated patients had lower rates of ICU admission (11% vs. 24%; *P* = 0.003) and ventilatory support (6% vs. 15%; *P* = 0.009). In unadjusted analyses, aspirin users had higher rates of cardiovascular complications (27% vs. 16%; *P* = 0.028) and lower 3-year survival (57% vs. 72%; *P* = 0.008). In contrast, NSAID users had lower rates of ICU admission (7% vs. 18%; *P* = 0.042), cardiovascular complications (4% vs. 23%; *P* = 0.001), and renal complications (9% vs. 25%; *P* = 0.010), and higher 1-year (98% vs. 78%; *P* = 0.0004) and 3-year survival (89% vs. 63%; *P* = 0.0001). After adjustment, aspirin use was not independently associated with any study outcome. Early NSAID use was independently associated with lower odds of renal complications (aOR, 0.35; 95% CI, 0.13–0.97; *P* = 0.044) and lower hazards of 1-year (aHR, 0.11; 95% CI, 0.01–0.81; *P* = 0.030) and 3-year mortality (aHR, 0.33; 95% CI, 0.14–0.77; *P* = 0.011). Sensitivity analyses using the Charlson Comorbidity Index yielded similar findings. Prospective studies are needed to determine whether early non-aspirin NSAID use improves clinical outcomes in adults hospitalized with influenza.

## Introduction

Seasonal influenza remains a major public health concern, causing substantial morbidity and mortality worldwide. The annual burden varies with circulating viral strains, their virulence, the timing and duration of influenza activity, vaccine effectiveness, vaccination coverage, and host immune responses. In the United States, seasonal influenza caused an estimated 9.3–41 million illnesses, 120,000–710,000 hospitalizations, and 6,300–52,000 deaths annually between 2010 and 2024 [1]. Globally, influenza causes an estimated 3–5 million cases of severe illness and up to 650,000 respiratory-related deaths each year [2].

An effective immune response is essential for viral clearance and recovery. However, excessive production of proinflammatory cytokines and chemokines can cause tissue damage, acute lung injury, and multi-organ dysfunction [3–6]. Influenza has also been linked to an increased risk of acute myocardial infarction, likely mediated by inflammation-induced platelet activation [7–9]. In addition, infection promotes platelet-endothelial adhesion in the lung via fibronectin, contributing to acute lung injury [10]. Influenza vaccination was associated with lower risk of cardiovascular events and all-cause mortality among individuals with high cardiovascular risk or established cardiovascular disease [11–13]. Antiviral therapy with neuraminidase inhibitors (NAIs) remains the standard of care and improves outcomes, however, complications and deaths continue to occur despite their use [14].

Given the central role of inflammation and platelet activation in influenza pathogenesis, adjunctive therapies targeting these pathways warrant investigation. Aspirin irreversibly inhibits thromboxane A_2_ synthesis, thereby inhibiting platelet aggregation and reducing cardiovascular risk, and has also demonstrated antiviral activity in preclinical influenza models [15]. Non-aspirin nonsteroidal anti-inflammatory drugs (NSAIDs), including selective cyclooxygenase-2 (COX-2) inhibitors, reduce inflammation by inhibiting the inducible COX-2 enzyme and have demonstrated antiviral activity against influenza A (H5N1) in vitro [16].

This study evaluated the associations of early aspirin and non-aspirin NSAID use with clinical outcomes in adults hospitalized with laboratory-confirmed influenza.

## Methods

### Subjects and study design

This retrospective cohort study evaluated the associations of influenza vaccination and early aspirin and non-aspirin nonsteroidal anti-inflammatory drug (NSAID) use with clinical outcomes in adults hospitalized with influenza. Eligible participants were adults aged 18 years or older admitted to the University of Minnesota Medical Center (UMMC) between 01/01/2016 and 31/12/2018. Early medication use was defined as medication administration before hospital presentation, on the day of presentation, or within 24 hours of hospital admission. Influenza was diagnosed using rapid molecular assays, including influenza/respiratory syncytial virus (RSV) polymerase chain reaction (PCR), multiplex respiratory panel PCR, or rapid influenza antigen testing. Cases were defined as hospitalized adults with laboratory-confirmed influenza. Electronic health data used for this study were accessed between 01/06/2020 and 31/12/2022.

### Statistical methods

Descriptive statistics were used to summarize the study population. Data distribution was assessed for normality using the Shapiro–Wilk test. Continuous variables, which were not normally distributed, were reported as medians with interquartile ranges (IQRs), and categorical variables were reported as counts and percentages.

Bivariate analyses were conducted to compare baseline characteristics and outcomes between groups. Continuous variables were analyzed using the Wilcoxon rank-sum test, and categorical variables were analyzed using the chi-square test or Fisher’s exact test when expected cell counts were sparse. Comparisons were made between vaccinated and unvaccinated patients, aspirin users and non-users, and NSAID users and non-users.

Multivariable analyses were performed to examine the associations of aspirin and NSAID use with clinical outcomes among adults hospitalized with laboratory-confirmed influenza. Logistic regression models were used to evaluate binary outcomes, including intensive care unit (ICU) admission, ventilatory support, cardiovascular complications, renal complications, and 30-day readmission. Cox proportional hazards regression models were used to evaluate time-to-event outcomes, including time to hospital discharge and time to death.

All models were adjusted for age, sex, race, body mass index (BMI), smoking status, influenza vaccination status, and cardiovascular burden. Cardiovascular burden was defined as the total number of baseline cardiovascular comorbidities present at hospital admission, including coronary artery disease, congestive heart failure, atrial fibrillation, hypertension, peripheral vascular disease, and cerebrovascular disease, with each condition contributing one point to the composite score. For the multivariable analyses, race was collapsed into two categories (White and all others) because of sparse data in the non-White racial groups. Sensitivity analyses were performed by replacing cardiovascular burden with the Charlson Comorbidity Index in the multivariable models [17]. An additional sensitivity analysis was conducted by restricting the exposure definition to preadmission aspirin and non-aspirin NSAID use.

All statistical analyses were conducted using SAS version 9.4 (SAS Institute Inc., Cary, NC). A *P* value < 0.05 was considered statistically significant. This was an observational study with a limited sample size. Therefore, the analyses were considered exploratory, and no adjustments were made for multiple testing.

## Results

### Baseline characteristics by influenza vaccination status

Among 2,816 hospitalized adults screened for influenza-like illness, 320 had laboratory-confirmed influenza. Of the 320 patients, 180 (56.3%) were vaccinated and 140 (43.7%) were unvaccinated. Vaccinated patients were significantly older than unvaccinated patients (median age, 64 (IQR, 49-74) vs. 58 (IQR, 35-71) years; *P* = 0.007). Sex, race, BMI, smoking status, and the prevalence of major comorbidities, including cardiovascular disease, respiratory disease, diabetes mellitus, renal disease, malignancy, and immunosuppression, were similar between vaccinated and unvaccinated patients.

Despite similar prevalences of individual comorbidities, vaccinated patients had a higher overall comorbidity burden than unvaccinated patients, as reflected by higher Charlson Comorbidity Index scores (median, 3 (IQR, 1-5) vs. 2 (IQR, 1-4); *P* =0.013) and cardiovascular burden scores (*P* =0.039).

Compared with unvaccinated patients, vaccinated patients were more likely to receive preadmission anticoagulants (20.6% vs. 7.9%; *P* = 0.002), antibiotics (47.8% vs. 30.7%; *P* =0.002), and vitamin D supplementation (52.8% vs. 34.3%; *P* = 0.001). Similar findings were observed when in-hospital medication use was included, with higher use of anticoagulants (25.6% vs. 10.7%; *P* = 0.001) and vitamin D supplementation (55.0% vs. 36.4%; *P* =0.001) among vaccinated patients. Use of aspirin, non-aspirin NSAIDs, acetaminophen, statins, corticosteroids, in-hospital antibiotics, and oseltamivir was similar between the groups (Table 1).

**Table 1.**
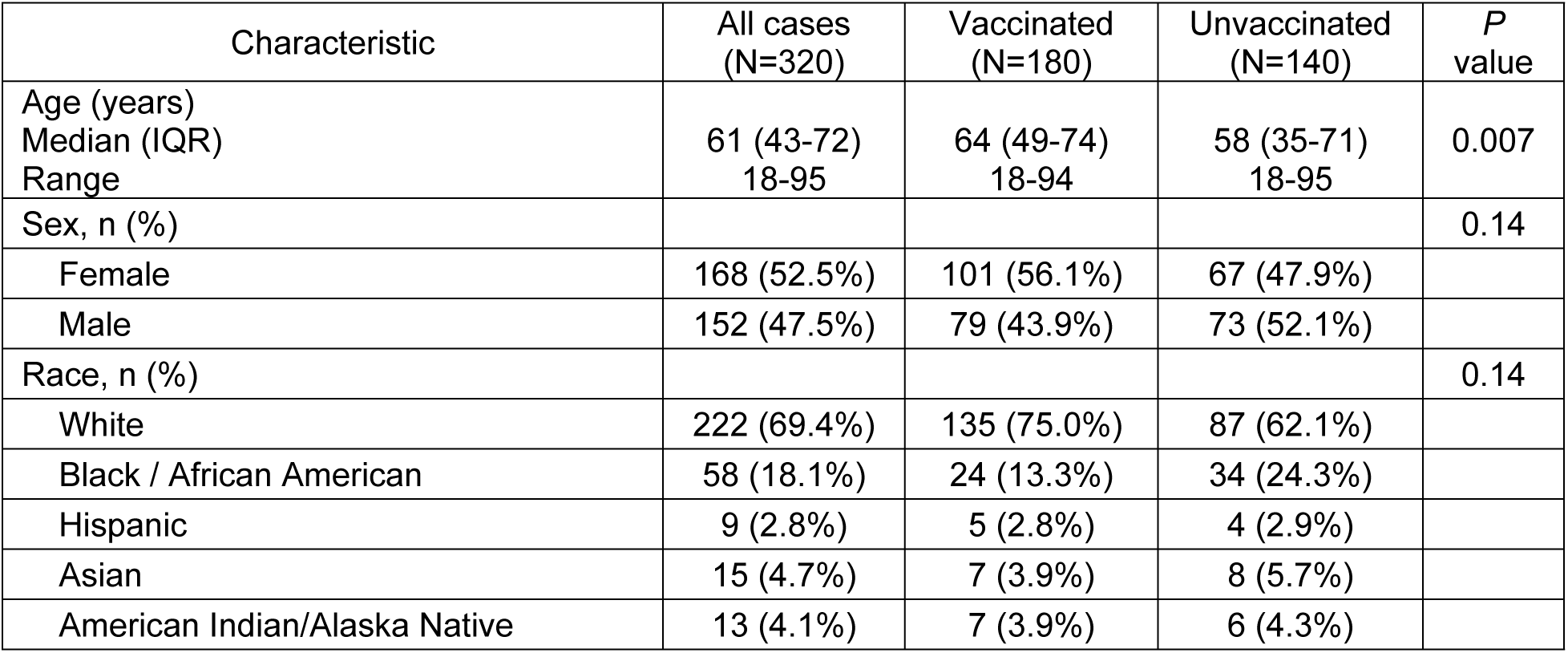
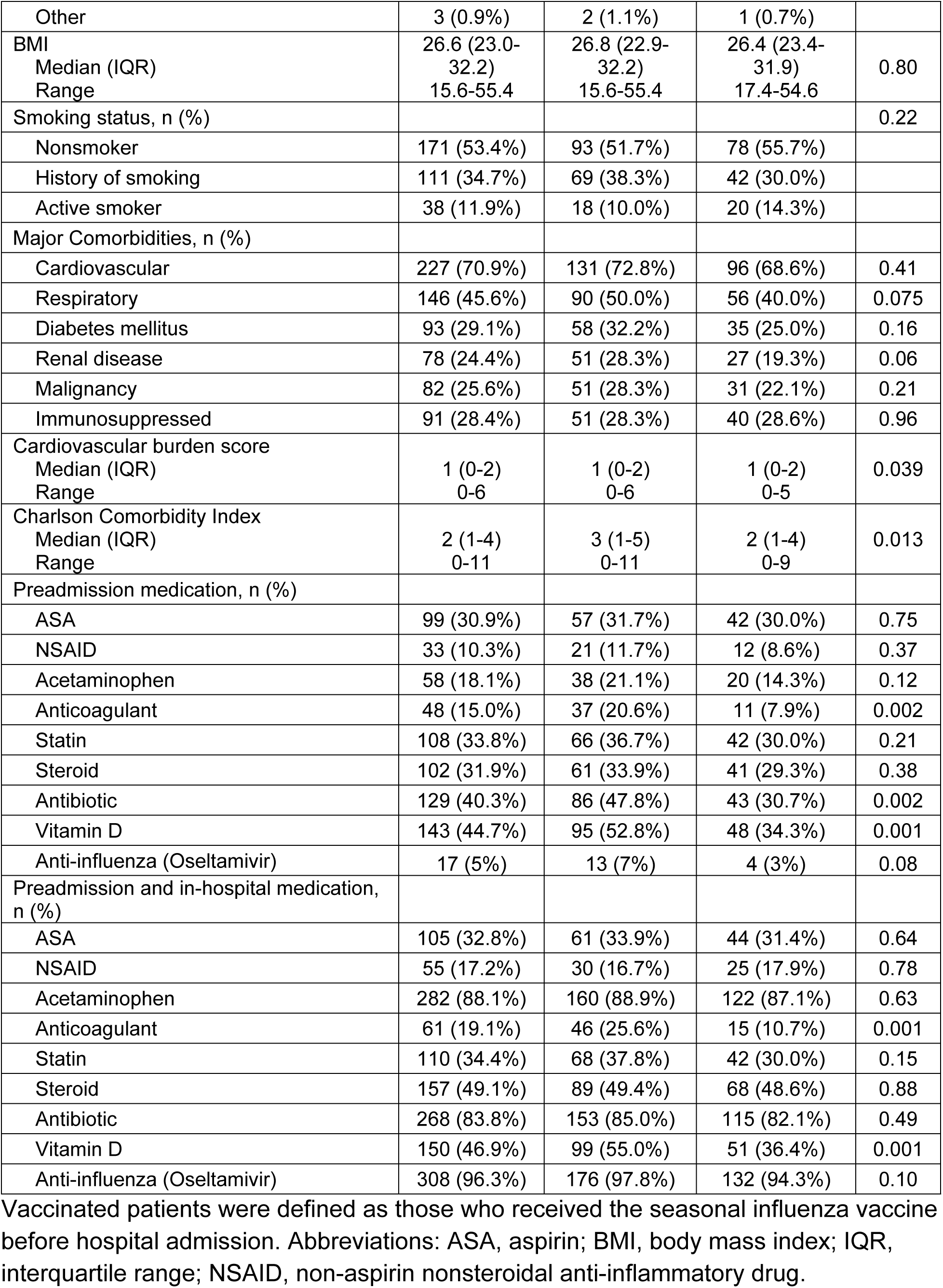
Baseline characteristics of adults hospitalized with influenza by influenza vaccination status.

### Clinical outcomes by influenza vaccination status

Compared with unvaccinated patients, vaccinated patients had significantly lower rates of ICU admission (11% vs. 24%; *P* = 0.003) and ventilatory support (6% vs. 15%; *P* = 0.009). No significant differences were observed in length of hospital stay, cardiovascular complications, renal complications, in-hospital mortality, or 30-day readmission. Although 1-year (78% vs. 85%; *P* = 0.13) and 3-year (63% vs. 73%; *P* = 0.057) survival rates were lower among vaccinated patients, these differences were not statistically significant (Table 2).

**Table 2.** Clinical outcomes of adults hospitalized with influenza by vaccination status.

| Outcome | All cases (N=320) | Vaccinated (N=180) | Unvaccinated (N=140) | *P* value |
| --- | --- | --- | --- | --- |
| Length of hospital stay (days), median (IQR) | 5 (3-9) | 5 (3-8) | 4 (3-9) | 0.38 |
| Intensive Care Unit (ICU) admission, n (%) | 53 (17%) | 20 (11%) | 33 (24%) | 0.003 |
| Required ventilatory support, n (%) | 32 (10%) | 11 (6%) | 21 (15%) | 0.009 |
| Cardiovascular complications, n (%) | 63 (20%) | 33 (18%) | 30 (21%) | 0.49 |
| Renal complications, n (%) | 71 (22%) | 37 (21%) | 34 (24%) | 0.43 |
| Died in hospital, n (%) | 15 (5%) | 7 (4%) | 8 (6%) | 0.44 |
| Readmission within 30 days, n (%) | 52 (16%) | 33 (18%) | 19 (14%) | 0.25 |
| 1-year survival, n (%) | 260 (81%) | 141 (78%) | 119 (85%) | 0.13 |
| 3-year survival, n (%) | 215 (67%) | 113 (63%) | 102 (73%) | 0.057 |

Vaccinated patients were defined as those who received the seasonal influenza vaccine before hospital admission. One-year and 3-year survival were calculated from the date of hospital admission for influenza. Abbreviations: ICU, intensive care unit; IQR, interquartile range.

### Baseline characteristics by early aspirin and non-aspirin NSAID use

Aspirin users were significantly older than non-users (median age, 70 (IQR, 61-77) vs. 55 (IQR, 34-67) years; *P* < 0.0001). Compared with non-users, aspirin users had a higher prevalence of cardiovascular disease (93% vs. 60%; *P* < 0.0001) and renal disease (34% vs. 20%; *P* = 0.004), as well as greater cardiovascular burden (median score, 2 (IQR, 1-3) vs. 1 (IQR, 0-2); *P* < 0.0001) and Charlson Comorbidity Index scores (median, 4 (IQR, 2-5) vs. 2 (IQR, 1-4); *P* < 0.0001). Aspirin users were more likely to have a history of smoking than non-users (47% vs. 29%; *P* = 0.002). Sex, race, BMI, influenza vaccination status, and the prevalence of respiratory disease did not differ significantly between the groups.

In contrast, NSAID users were significantly younger than non-users (median age, 47 (IQR, 31–67) vs. 63 (IQR, 47–73) years; *P* = 0.0002) and were more likely to be female (65% vs. 50%; *P* = 0.035). Compared with non-users, NSAID users had a lower prevalence of cardiovascular disease (45% vs. 76%; *P* < 0.0001), and renal disease (4% vs. 29%; *P* < 0.0001), as well as lower cardiovascular burden scores (median, 0 (IQR, 0–1) vs. 1 (IQR, 0–2); *P* < 0.0001) and Charlson Comorbidity Index scores (median, 1 (IQR, 0–3) vs. 3 (IQR, 1–5); *P* < 0.0001). Race, BMI, smoking status, influenza vaccination status, and the prevalence of respiratory disease did not differ significantly between the groups (Table 3).

**Table 3.**
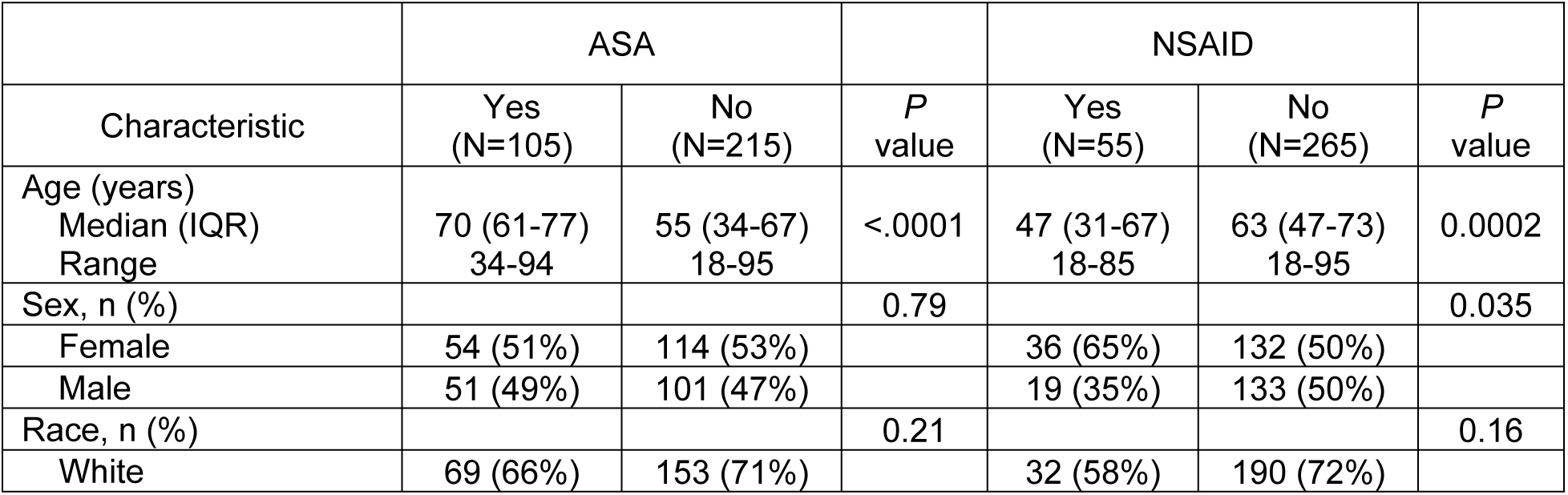
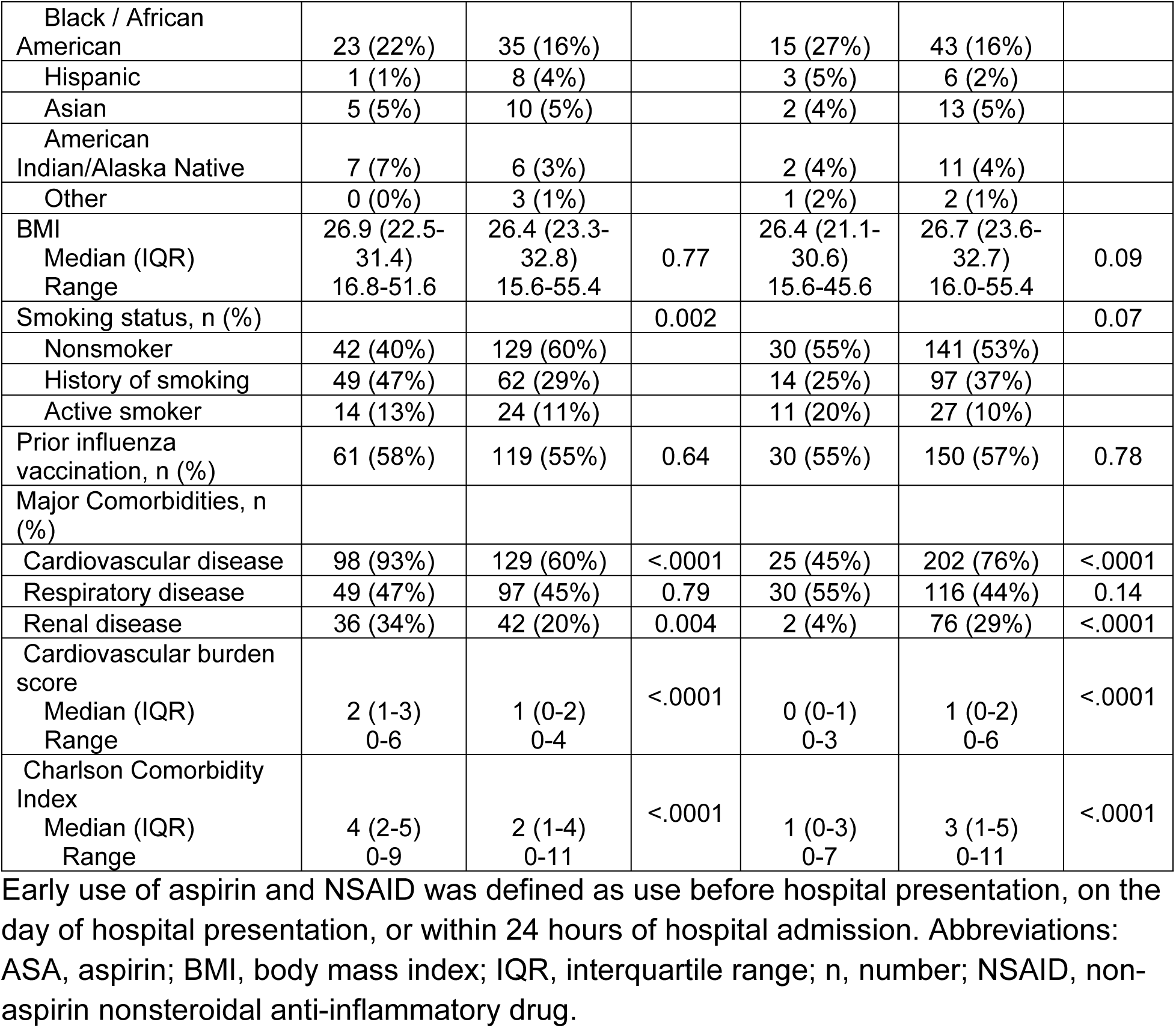
Baseline characteristics of adults hospitalized with influenza by early aspirin and non-aspirin NSAID use.

### Clinical outcomes by early aspirin and non-aspirin NSAID use

Compared with non-users, aspirin users had higher rates of cardiovascular complications (27% vs. 16%; *P* = 0.028) and lower 3-year survival (57% vs. 72%; *P* = 0.008), whereas no significant differences were observed for the remaining clinical outcomes. In contrast, NSAID users had significantly lower rates of ICU admission (7% vs. 18%; *P* = 0.042), cardiovascular complications (4% vs. 23%; *P* = 0.001), and renal complications (9% vs. 25%; *P* = 0.010), as well as higher 1-year survival (98% vs. 78%; *P* = 0.0004) and 3-year survival (89% vs. 63%; *P* = 0.0001) than non-users. Length of hospital stay, ventilatory support, in-hospital mortality, and 30-day readmission did not differ significantly between NSAID users and non-users (Table 4).

**Table 4.** Clinical outcomes of adults hospitalized with influenza by early aspirin and non-aspirin NSAID use.

| Outcome | ASA |  | *P* value | NSAID |  | *P* value |
| --- | --- | --- | --- | --- | --- | --- |
|  | Yes (N=105) | No (N=215) |  | Yes (N=55) | No (N=265) |  |
| Length of hospital stay (days), median (IQR) | 4 (3-8) | 5 (3-9) | 0.87 | 4 (3-6) | 5 (3-9) | 0.14 |
| Intensive Care Unit (ICU) admission, n (%) | 15 (14%) | 38 (18%) | 0.44 | 4 (7%) | 49 (18%) | 0.042 |
| Required ventilatory support, n (%) | 8 (8%) | 24 (11%) | 0.32 | 3 (5%) | 29 (11%) | 0.22 |
| Cardiovascular complications, n (%) | 28 (27%) | 35 (16%) | 0.028 | 2 (4%) | 61 (23%) | 0.001 |
| Renal complications, n (%) | 29 (28%) | 42 (20%) | 0.10 | 5 (9%) | 66 (25%) | 0.010 |
| Died in hospital, n (%) | 6 (6%) | 9 (4%) | 0.54 | 1 (2%) | 14 (5%) | 0.27 |
| Readmission within 30 days, n (%) | 18 (17%) | 34 (16%) | 0.76 | 9 (16%) | 43 (16%) | 0.98 |
| 1-year survival, n (%) | 83 (79%) | 177 (82%) | 0.48 | 54 (98%) | 206 (78%) | 0.0004 |
| 3-year survival, n (%) | 60 (57%) | 155 (72%) | 0.008 | 49 (89%) | 166 (63%) | 0.0001 |

Early use of aspirin and NSAID was defined as use before hospital presentation, on the day of hospital presentation, or within 24 hours of hospital admission. One-year and 3-year survival were calculated from the date of hospital admission for influenza. Abbreviations: ASA, aspirin; ICU, intensive care unit; IQR, interquartile range; NSAID, non-aspirin nonsteroidal anti-inflammatory drug.

### Multivariable analyses

Given the substantial differences in age and comorbidity burden between aspirin and NSAID users, multivariable analyses were performed to determine whether the observed associations persisted after adjustment for potential confounders. After adjustment for age, sex, race, smoking status, influenza vaccination status, and cardiovascular burden, aspirin use was not independently associated with any study outcome. In contrast, early NSAID use was independently associated with lower odds of renal complications (aOR, 0.35; 95% CI, 0.13–0.97; *P* = 0.044) and lower hazards of 1-year mortality (aHR, 0.11; 95% CI, 0.01–0.81; *P* = 0.030) and 3-year mortality (aHR, 0.33; 95% CI, 0.14–0.77; *P* = 0.011). Lower odds of ICU admission and cardiovascular complications were also observed but did not reach statistical significance. No significant associations were observed for ventilatory support, 30-day readmission, or length of hospital stay (Table 5).

**Table 5.** Multivariable-adjusted associations of early aspirin and non-aspirin NSAID use with clinical outcomes.

| Outcome | ASA |  | NSAID |  |
| --- | --- | --- | --- | --- |
|  | Adjusted effect estimates (95% CI) | *P* value | Adjusted effect estimates (95% CI) | *P* value |
|  | aOR (95% CI) |  | aOR (95% CI) |  |
| Intensive Care Unit (ICU) admission | 0.66 (0.30,1.47) | 0.31 | 0.33 (0.11,1.01) | 0.051 |
| Required ventilator support | 0.57 (0.20,1.62) | 0.29 | 0.44 (0.12,1.65) | 0.23 |
| Cardiovascular complications | 0.97 (0.49,1.93) | 0.93 | 0.24 (0.06,1.07) | 0.06 |
| Renal complications | 1.05 (0.54,2.03) | 0.89 | 0.35 (0.13,0.97) | 0.044 |
| Readmission within 30 days | 1.82 (0.84,3.96) | 0.13 | 0.77 (0.33,1.78) | 0.54 |
|  | aHR (95% CI) |  | aHR (95% CI) |  |
| Length of hospital stay (time to discharge) | 1.10 (0.82,1.48) | 0.52 | 1.26 (0.92,1.72) | 0.15 |
| 1-year mortality | 0.67 (0.37,1.21) | 0.18 | 0.11 (0.01,0.81) | 0.030 |
| 3-year mortality | 0.90 (0.58,1.40) | 0.64 | 0.33 (0.14,0.77) | 0.011 |

Early use of aspirin and NSAID was defined as use before hospital presentation, on the day of hospital presentation, or within 24 hours of hospital admission. One-year and 3-year mortality were calculated from the date of hospital admission for influenza. Adjusted for age, sex, race, BMI, smoking status, influenza vaccination status, and cardiovascular burden. Abbreviations: aOR, adjusted odds ratio; aHR, adjusted hazard ratio; ASA, aspirin; CI, confidence interval; ICU, intensive care unit; NSAID, non-aspirin nonsteroidal anti-inflammatory drug.

### Sensitivity analyses

Sensitivity analyses replacing cardiovascular burden with the Charlson Comorbidity Index yielded findings that were generally consistent with the primary analyses. Aspirin use remained unassociated with the study outcomes, whereas early NSAID use remained associated with lower odds of renal complications and lower hazards of 1-year and 3-year mortality. In addition, lower odds of ICU admission and cardiovascular complications among NSAID users reached statistical significance (S1 Table). When the exposure definition was restricted to preadmission aspirin and NSAID use, the direction of the associations remained consistent, although none of the associations reached statistical significance (S2 Table).

## Discussion

In this retrospective cohort study of adults hospitalized with influenza, influenza vaccination was associated with lower rates of ICU admission and ventilatory support. Early aspirin use was associated with less favorable outcomes in unadjusted analyses, but these associations were no longer statistically significant after adjustment for baseline differences. In contrast, early NSAID use was independently associated with lower risks of renal complications and reduced 1- and 3-year mortality after multivariable adjustment. Lower risks of ICU admission and cardiovascular complications were also observed, but these associations were no longer statistically significant after adjustment. Sensitivity analyses yielded generally consistent findings, supporting the robustness of the primary analyses.

In our study, vaccinated patients were less likely to require ICU admission or ventilatory support. Although vaccinated patients also showed a trend toward improved long-term survival, the difference was not statistically significant. These findings are consistent with previous evidence that influenza vaccination reduces the severity of influenza illness. A systematic review and meta-analysis found that influenza vaccination reduced the risk of severe influenza requiring hospitalization [18]. In addition, a CDC analysis estimated that influenza vaccination prevented substantial numbers of influenza-associated hospitalizations and deaths during the 2017–2018 influenza season [19].

Influenza virus induces both direct cytopathic injury and a dysregulated host inflammatory response that contributes to pulmonary damage and systemic complications [3, 20–22]. Modulation of this inflammatory response may therefore influence disease severity and clinical outcomes. Non-aspirin NSAIDs inhibit cyclooxygenase (COX) enzymes, thereby reducing prostaglandin synthesis and attenuating the inflammatory response, providing a plausible mechanism for their potential effects during influenza infection (23).

Historically, clinicians have been cautious about NSAID use during acute respiratory infections because of concerns that NSAIDs may mask symptoms, delay diagnosis and appropriate treatment, and potentially increase the risk of severe clinical complications [24]. However, evidence regarding NSAID use in influenza remains limited. A nationwide cohort study of adults hospitalized with influenza found that NSAID use was not associated with ICU admission or 30-day mortality in adjusted analyses. Based on these findings, the authors concluded that their results did not support strong recommendations against NSAID use in patients with viral pneumonia. The study also reported similar associations in patients with bacterial pneumonia and an increased risk of pleuropulmonary complications among NSAID users in that cohort [25]. In another retrospective study of critically ill patients with pandemic H1N1 influenza found that preadmission NSAID use was not independently associated with mortality after multivariable adjustment [26]. In contrast, early NSAID use in our study remained independently associated with lower risks of renal complications and reduced 1- and 3-year mortality in multivariable analyses. Early NSAID use was also associated with lower adjusted risks of ICU admission and cardiovascular complications, although these associations were not statistically significant. Several methodological differences may explain these findings, including the evaluation of early non-aspirin NSAID exposure, the separate assessment of aspirin and non-aspirin NSAIDs, adjustment for important baseline characteristics, and the inclusion of long-term mortality and influenza-associated cardiovascular and renal complications as study outcomes.

Concerns regarding NSAID-associated nephrotoxicity have limited their use during acute illness. However, in our study, early NSAID use was independently associated with a lower risk of renal complications and was not associated with an increased risk of acute renal injury. These findings are consistent with previous reports suggesting that, in stable and adequately hydrated individuals, NSAIDs generally carry a low risk of nephrotoxicity [27]. NSAID-associated renal injury is more likely to occur in the setting of dehydration, underlying comorbidities, or prolonged use. Ibuprofen and ketorolac were the most frequently administered NSAIDs in our cohort, whereas celecoxib and indomethacin were used infrequently. The limited use of NSAIDs other than ibuprofen and ketorolac precluded meaningful comparisons of individual NSAID-specific effects, highlighting the need for future studies evaluating individual agents.

Although this observational study cannot establish causality, the consistent associations observed with early NSAID use suggest that earlier modulation of the host inflammatory response may be associated with fewer influenza-related complications and improved long-term survival. The consistency of these findings across sensitivity analyses further supports the robustness of the observed associations. These findings warrant confirmation in prospective studies to determine whether early NSAID use improves clinical outcomes in adults hospitalized with influenza.

In contrast to the findings for NSAID use, aspirin was not independently associated with clinical outcomes after multivariable adjustment. The unfavorable unadjusted associations observed among aspirin users may have been explained, at least in part, by their older age and greater cardiovascular burden because these associations were no longer evident after multivariable adjustment. Aspirin irreversibly inhibits cyclooxygenase-1 (COX-1), thereby suppressing thromboxane A₂ synthesis and producing sustained inhibition of platelet aggregation, providing a rationale for reducing influenza-associated thrombo-inflammatory complications [28]. Despite this rationale, clinical evidence evaluating aspirin specifically in influenza remains limited. Consistent with our findings, an exploratory analysis of critically ill adults with pandemic H1N1 influenza found that preadmission aspirin use was not independently associated with mortality after adjustment for potential confounders [26].

Sensitivity analyses supported the robustness of the primary findings. Similar associations were observed when the Charlson Comorbidity Index was substituted for the cardiovascular burden score in the sensitivity analyses. When the exposure definition was restricted to preadmission medication use only, the associations remained in the same direction but no longer reached statistical significance, likely reflecting reduced statistical power. These findings suggest that the observed associations are robust across different analytic approaches and support further investigation into whether the timing of early non-aspirin NSAID initiation during influenza infection influences clinical outcomes.

This study has several limitations. First, as a single-center retrospective cohort study with a relatively small sample size, it is subject to residual confounding and selection bias. Because treatment allocation was not randomized, the indications for aspirin and NSAID use may have differed between patients, and residual confounding by indication may have persisted despite multivariable adjustment and sensitivity analyses. Second, information on outpatient medication adherence was unavailable, and the timing, dosage, and duration of aspirin and NSAID use could not be reliably determined. Third, the small number of patients receiving less commonly used NSAIDs precluded meaningful subgroup analyses. Finally, because of the observational nature of this study, the findings do not establish causality and require confirmation in prospective studies.

### Conclusion

In this study, influenza vaccination was associated with lower rates of ICU admission and ventilatory support. These findings are consistent with previous studies and reinforce the role of influenza vaccination as the cornerstone of influenza prevention. However, severe complications and deaths continue to occur despite vaccination and antiviral therapy, highlighting the need for adjunctive strategies to further improve clinical outcomes. In this study, early non-aspirin NSAID use was independently associated with lower risks of renal complications and reduced 1- and 3-year mortality, whereas early aspirin use was not independently associated with clinical outcomes after multivariable adjustment. These findings emphasize the importance of distinguishing aspirin from non-aspirin NSAIDs in influenza research and raise the possibility that earlier initiation of non-aspirin NSAIDs during influenza infection may be associated with improved clinical outcomes. Prospective studies are warranted to determine whether these observed associations are causal and to define the optimal timing, dosage and role of non-aspirin NSAIDs as adjunctive therapy in adults hospitalized with influenza.

## Data Availability

All relevant data are within the paper and its Supporting Information files.

## Acknowledgements

We thank the staff at the Clinical Research Support Center for their assistance during the early stages of this research.

## Author Contributions

**Conceptualization:** Suk Yin Chan-Colenbrander

**Data curation:** Suk Yin Chan-Colenbrander

**Formal analysis:** Suk Yin Chan-Colenbrander, Qi Wang

**Funding acquisition:** Suk Yin Chan-Colenbrander

**Investigation:** Suk Yin Chan-Colenbrander

**Methodology:** Suk Yin Chan-Colenbrander, Qi Wang

**Supervision:** Suk Yin Chan-Colenbrander

**Visualization:** Suk Yin Chan-Colenbrander, Qi Wang

**Writing - original draft:** Suk Yin Chan-Colenbrander

**Writing – review & editing:** Suk Yin Chan-Colenbrander, Qi Wang

## Supporting Information

**S1 Table.** Sensitivity analysis of multivariable-adjusted associations of early aspirin and non-aspirin NSAID use with clinical outcomes using the Charlson Comorbidity Index.

**S2 Table.** Sensitivity analysis of multivariable-adjusted associations of preadmission aspirin and non-aspirin NSAID use with clinical outcomes.

**S1 Data.** De-identified data for adults hospitalized with laboratory-confirmed influenza.

